# What level of neutralising antibody protects from COVID-19?

**DOI:** 10.1101/2021.03.09.21252641

**Authors:** David S Khoury, Deborah Cromer, Arnold Reynaldi, Timothy E Schlub, Adam K Wheatley, Jennifer A Juno, Kanta Subbarao, Stephen J Kent, James A Triccas, Miles P Davenport

**Affiliations:** Kirby Institute, University of New South Wales, Sydney, Australia; Sydney School of Public Health, Faculty of Medicine and Health, University of Sydney, Sydney, New South Wales, Australia; Department of Microbiology and Immunology, University of Melbourne at the Peter Doherty Institute for Infection and Immunity, Melbourne, Australia; WHO Collaborating Centre for Reference and Research on Influenza, The Peter Doherty Institute for Infection and Immunity, Melbourne, Australia; Australian Research Council Centre for Excellence in Convergent Bio-Nano Science and Technology, University of Melbourne, Melbourne, Victoria, Australia; Melbourne Sexual Health Centre and Department of Infectious Diseases, Alfred Hospital and Central Clinical School, Monash University, Melbourne, Victoria, Australia; School of Medical Sciences and Marie Bashir Institute, Faculty of Medicine and Health, The University of Sydney, Camperdown, NSW, Australia

## Abstract

Both previous infection and vaccination have been shown to provide potent protection from COVID-19. However, there are concerns that waning immunity and viral variation may lead to a loss of protection over time. Predictive models of immune protection are urgently needed to identify immune correlates of protection to assist in the future deployment of vaccines. To address this, we modelled the relationship between in vitro neutralisation levels and observed protection from SARS-CoV-2 infection using data from seven current vaccines as well as convalescent cohorts. Here we show that neutralisation level is highly predictive of immune protection. The 50% protective neutralisation level was estimated to be approximately 20% of the average convalescent level (95% CI = 14-28%). The estimated neutralisation level required for 50% protection from severe infection was significantly lower (3% of the mean convalescent level (CI = 0.7-13%, p = 0.0004). Given the relationship between in vitro neutralization titer and protection, we then used this to investigate how waning immunity and antigenic variation might affect vaccine efficacy. We found that the decay of neutralising titre in vaccinated subjects over the first 3-4 months after vaccination was at least as rapid as the decay observed in convalescent subjects. Modelling the decay of neutralisation titre over the first 250 days after immunisation predicts a significant loss in protection from SARS-CoV-2 infection will occur, although protection from severe disease should be largely retained. Neutralisation titres against some SARS-CoV-2 variants of concern are reduced compared to the vaccine strain and our model predicts the relationship between neutralisation and efficacy against viral variants. Our analyses provide an evidence-based prediction of SARS-CoV-2 immune protection that will assist in developing vaccine strategies to control the future trajectory of the pandemic.

## Main text

SARS-CoV-2 has spread globally over the last year, infecting an immunologically naïve population and causing significant morbidity and mortality. Immunity to SARS-CoV-2 induced either through natural infection or vaccination has been shown to afford a degree of protection and/or reduce the risk of clinically significant outcomes. For example, seropositive recovered subjects have been estimated to have 89% protection from reinfection ^1^, and vaccine efficacies from 50 to 95% have been reported ^2^. However, the duration of protective immunity is unclear, with primary immune responses inevitably waning ^3-5^, and ongoing transmission of increasingly concerning viral variants that escape immune control ^6^.

A critical question at present is to identify the immune correlate(s) of protection from SARS-CoV-2 infection and therefore predict how changes in immunity will be reflected in clinical outcomes. A defined correlate of protection will permit both confidence in opening up economies and facilitate rapid improvements in vaccines and immunotherapies. In influenza infection, for example, a hemagglutination inhibition (HAI) titre of 1:40 is thought to provide 50% protection from influenza infection ^7^ (although estimates range from 1:17 – 1:110 ^8,9^). This level was established over many years using data from a standardised HAI assay^10^ applied to serological samples from human challenge and cohort studies. At present, however, there are few standardised assays for assessing SARS-CoV-2 immunity, little data comparing immune levels in susceptible versus resistant individuals, and no human challenge model ^11^.

The data currently available for SARS-CoV-2 infection includes immunogenicity data from phase 1 / 2 studies of vaccines and data on protection from preliminary reports from phase 3 studies and in seropositive convalescent individuals (see Supplementary Tables 1 and 2). While antiviral T and B cell memory certainly contribute some degree of protection, strong evidence of a protective role for neutralising serum antibodies exists. For example, passive transfer of neutralising antibodies can prevent severe SARS-CoV-2 infection in multiple animal models ^12,13^ and Regeneron has recently reported similar data in humans ^14^. We therefore focus our studies on *in vitro* virus neutralisation titres reported in studies of vaccinated and convalescent cohorts. Unfortunately, the phase 1 / 2 studies all use different assays for measuring neutralisation. Normalisation of responses against a convalescent serum standard has been suggested to provide greater comparability between the results from different assays^15^. Although all studies compare immune responses after vaccination against the responses in convalescent individuals, the definition of convalescence is not standardised across studies. Similarly, in phase 3 studies the timeframes of study and case-definitions of infection also vary amongst studies (see Supplementary Table 2). Recognising these limitations, we aimed to investigate the relationship between vaccine immunogenicity and protection.

To compare neutralisation titres across studies we determined the mean and standard deviation (on log-scale) of the neutralisation titre in each study. These were normalised to the mean convalescent titre using the same assay in the same study (noting that the definition of convalescence was also not standardised across studies and a variable number of convalescent samples are studied). We then compared this normalised neutralisation level in each study against the corresponding protective efficacy reported from the phase 3 clinical trials. Despite the known inconsistencies between studies, comparison of normalised neutralisation levels and vaccine efficacy demonstrates a remarkably strong non-linear relationship between average neutralization level and reported protection across different vaccines (Spearman r=0.905; p=0.0046, Figure 1A).

**Figure 1:**
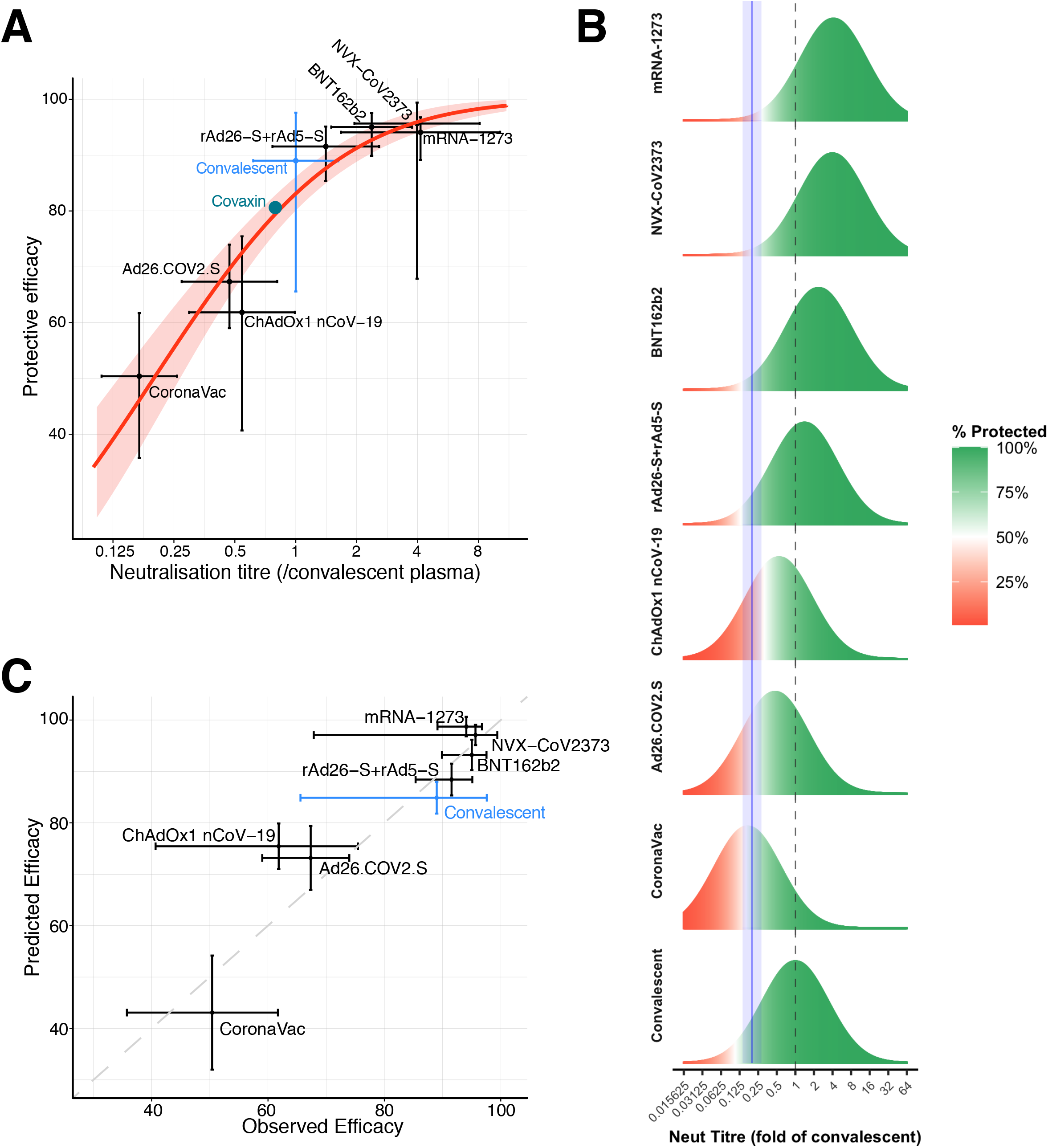
Understanding the relationship between neutralisation and protection. A) Relationship between neutralisation level and protection from SARS-CoV-2 infection. The reported mean neutralisation level (x-axis) from phase 1 / 2 trials and protective efficacy from phase 3 trials (y-axis) for seven vaccines, as well as the protection observed in a seropositive convalescent cohort are shown (details of data sources in Supplementary Tables 1 and 2). Mean / point estimates are indicated by dots (95% confidence intervals are indicated as whiskers). Red solid line indicates the best fit of the logistic model and shaded red indicates the 95% predictive interval of the model. The mean neutralisation level and protective efficacy of the Covaxin vaccine is indicated as a green circle (data from this study was only available after modelling was complete and did not contribute to fitting). B) Schematic illustration of the logistic approach to identifying the protective neutralisation level. The data for each study includes the mean and distribution of the in vitro neutralisation titre against SARS-CoV-2 in vaccinated or convalescent subjects (as a proportion of the titre in convalescent subjects)(green/red bell curve), accompanied by a level of protective efficacy for the same regimen. The efficacy is illustrated by the proportion of bell curve ‘protected’ (green) versus ‘susceptible (red) for individual studies (shaded areas in between reflect the changing risk). The modelling fits the optimal 50% protective neutralisation level (blue dashed line, shaded areas indicate 95% confidence limits) that best estimates the correct levels of protection observed across the different studies. C) Predictions of the ‘leave one out’ analysis. Modelling was repeated multiple times using all potential sets of seven vaccination / convalescent studies to predict the efficacy of the eighth study. Dots indicate point estimates and whiskers indicate 95% confidence / predictive intervals.

To further dissect the relationship between immunogenicity and protection in SARS-CoV-2 we considered the parallels with previous approaches to estimating a ‘50% protective titre’ in influenza infection. These historic studies in influenza involved comparison of HAI titres in infected versus uninfected subjects (in either natural infection or human challenge studies), and used logistic or receiver-operating characteristics (ROC) approaches to identify an HAI titre that provided protection ^7-9,16,17^). We adapted these approaches to analyse the existing data on ‘average neutralisation level’ in different studies and the observed level of protection from infection (details of statistical methods are provided in the Supplementary material).

We first fitted a logistic model to estimate the ‘50% protective neutralisation level’ (across all studies) that best predicted the protective effect observed in each study (consistent with the use of a logistic function to model protection in influenza serological studies ^16,17^). We estimate from this model a 50% protective neutralisation level of 19.9% (95% CI = 14.1% – 28.1%) of the mean convalescent level (Figures 1A and 1B), and that this model provided a good explanation of the relationship between average neutralisation level and protection across the studies. Since the model is dependent on the mean and distribution of neutralisation levels, we also estimated these using different approaches, leading to similar estimates (Fig. S2 and Supplementary Methods).

To relax the assumption that neutralisation levels are normally distributed in the above model, we also estimated the protective level using a distribution-free approach applied to the raw data for individual neutralisation levels reported in the studies. This method finds a protective neutralisation threshold that maximises the chances of classifying individuals as protected or not protected based on their neutralisation level being above or below the threshold and on the observed protective efficacy in Phase 3 trials (i.e.: it does not rely on neutralisation levels being normally distributed). We refer to this as the ‘protective neutralisation classification model’. Using this classification approach the estimated protective threshold was 28.6% (CI= 19.2% – 29.2%) of the average convalescent level. As expected, the estimated protective level using the classification method was slightly higher than the 50% protective level estimated using the logistic method, as the classification method essentially estimates a level of 100% protection instead of 50% protection.

This analysis suggests that the 50% neutralisation level for SARS-CoV-2 is approximately 20% of the average convalescent titre. This analysis shows that the relationship between the average protective levels and the observed protective efficacy across different vaccines can be predicted based on consideration of the level and distribution of neutralisation titres. To test the potential utility of this in predicting the protective efficacy of an unknown vaccine, we repeated our analysis with a ‘leave-one-out’ approach. That is, we fitted all possible groups of seven vaccine or convalescent studies and used this to predict the efficacy of the 8th. Figure 1C shows the results of using the logistic model of protection to predict the efficacy of each vaccine from the results of the other seven. In addition, after fitting the model to the data for eight vaccine / convalescent studies, the phase 3 results of another vaccine were released in a press release on 3 March 2021. Using the observed neutralisation level (a mean of 79.2% of the convalescent titre in that study (See Supplementary Table 1) the predicted efficacy of the new vaccine was 79.4%, (95% Predictive Interval: 76.0%-82.8%), which is in very close agreement with the reported efficacy of 80.6% ^18^, and suggests good predictive value of the model (Figure 1A).

As discussed above, a major caveat of our estimate of the relative protective level of antibodies in SARS-CoV-2 infection is that it includes aggregation of data collected from diverse neutralisation assays and clinical trial designs. Clearly, a more standardised approach to assays and trials design would allow refinement of these analyses^11^, although this has not occurred so far. In addition, the association of neutralisation with protection across these studies does not prove that neutralising antibodies are mechanistic in mediating protection. It is possible that neutralisation is correlated with other immune responses, leading to an apparent association (as has been suggested for the use of HAI titre in influenza ^19-21^). Another important refinement of this approach would be to have standardised measures of other serological and cellular responses to infection to identify if any of these provide a better predictive value than neutralisation. However, despite these limitations it is tempting to consider the implications of this protective titre for immunity to SARS-CoV-2 infection.

Recent studies have identified a decline in neutralisation titre with time after both natural infection and vaccination^3,4^. An average half-life of 90 days over the first 8 months of infection has been reported^5^ although our analysis suggests that an early rapid decline slows with time^3^. A major question is whether vaccine-induced responses may be more durable than those following infection. Limited studies have analysed the trajectory of neutralisation titre after vaccination^22^. To compare decay in neutralisation titre we fitted a model of exponential decay over equivalent time periods in data from either convalescent^3^ or mRNA vaccination^22^ cohorts. Comparing neutralisation titres measured between 26 and 115 days after either mRNA-based vaccination^22^ or symptom onset for post-infection sera ^3^, we found similar half-lives (65 days vs 58 days, respectively, p = 0.88, likelihood ratio test. See supplementary Figure S3). Although this comparison relies on limited data, it suggests that decay of vaccine-induced neutralisation is similar to that observed after natural infection.

If the relationship between neutralisation level and protection that we observe cross-sectionally between different vaccines is maintained over time, we can use our model to predict how the observed waning of neutralisation titres might affect vaccine effectiveness. Important caveats to such an extrapolation are that (i) it assumes that neutralisation is a major mechanism of protection (or that the mechanism of protection remains correlated with neutralisation), although B cell memory and T cell responses may be more durable ^3-5,23^ (and indeed B cell responses have been shown to increase following infection ^3^), (ii) it applies the decay of neutralisation observed in convalescence to the vaccine data, and (iii) it assumes that the decay in titre is the same regardless of the initial starting titre (whereas others have suggested faster decay for higher initial levels ^24^). These limitations notwithstanding, we used the estimated half-life of neutralisation titre of 90 days derived from a study of convalescent individuals ^5^ and modelled the decay of neutralisation and protection over the first 250 days after vaccination (Figure 2A). Our model predicts that waning neutralisation titre will have non-linear effects on protection from SARS-CoV-2 infection, depending on initial vaccine efficacy. For example, a vaccine starting with an initial efficacy of 95% would be expected to maintain 58% efficacy by 250 days. However, a response starting with an initial efficacy of 70% would be predicted to drop to 18% efficacy after 250 days. This analysis can also be used to estimate how long it would take a response of a given initial efficacy to drop to 50% (or 70%) efficacy, which may be useful in predicting the time until boosting is required to maintain a minimal level of efficacy (Figure 2B). Clearly data generated from standardised assays are needed to track the long-term decay of post-vaccination immune responses and their relationship to clinical protection. However, this model provides a framework that can be adapted to predict outcomes as further immune and protection data becomes available. Indeed, if a disconnect between the decay of neutralisation titre and protection is observed, this may be a direct pointer to the role of non-neutralising responses in protection.

**Figure 2:**
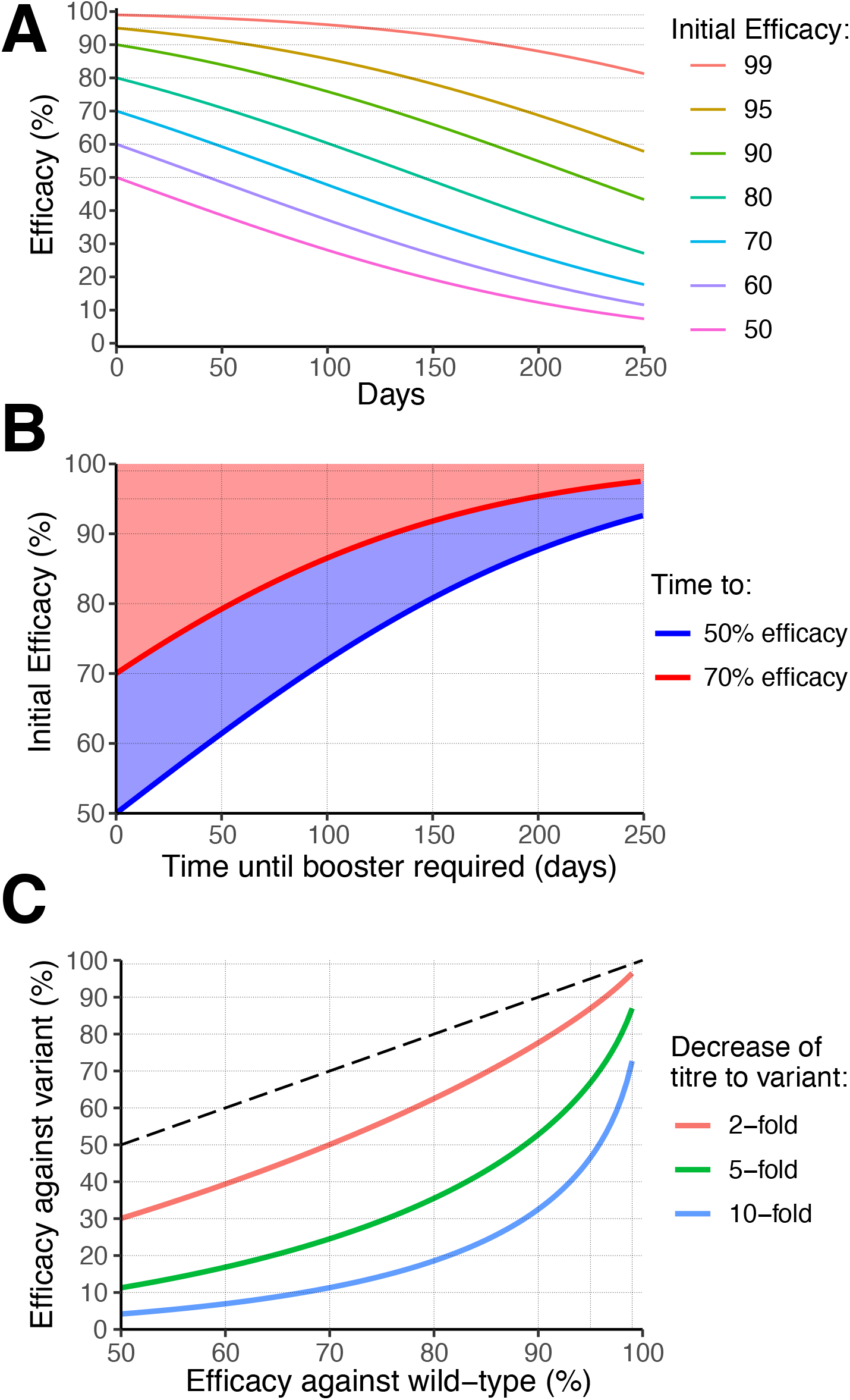
The effects of waning neutralisation titre on protection. A) Predicting the effects of declining neutralisation titre. Assuming the observed relationship between neutralisation level and protection is consistent over time, we estimate the decline in efficacy for vaccines starting with different levels of early protection. The model assumes a half-life of neutralisation titre of 90 days over the first 250 days (as observed in a convalescent cohort^5^). Coloured lines indicate the predicted trajectory for groups starting with different initial efficacy. B) Modelling the time for efficacy to drop to 70% (red line) or 50% (blue line) for scenarios with different initial efficacy (indicated on the y-axis). For example, for a group starting with an initial protective efficacy of 90% the model predicts that 70% efficacy will be reached after 131 days, and 50% efficacy after 221 days. C) Estimating the impact of viral antigenic variation on vaccine efficacy. *In vitro* studies have shown that neutralisation titres against some SARS-CoV-2 variants are reduced compared to titres against wild-type virus. If the relationship between neutralisation and protection remains constant, we can predict the difference in protective efficacy against wild-type and variant viruses from the difference in neutralisation level. For cohorts with a given initial protective efficacy measured against wild-type (vaccine strain) virus (x-axis), we model the impact of a two-fold (red), 5-fold (green) or 10-fold (blue) reduction in neutralisation titre to a variant virus. The y-axis indicates the predicted new efficacy against the variant strain. Dashed line indicates equal protection against wild-type and variant strains. Details of the data and modelling are provided in the Supplementary Material.

In addition to the effect of declining neutralisation titre over time, reduced neutralisation titres and reduced vaccine efficacy to different viral variants have also been observed ^6,25-28^. For example, it has been reported that the neutralisation titre against the B.1.351 variant in vaccinated individuals is between 7.6-fold and 9-fold lower compared to the early Victoria variant ^29^. Our model predicts that a lower neutralisation titre against a variant of concern will have a larger effect on vaccines for which protective efficacy against the wild-type virus was lower (Figure 2C). For example, a five-fold lower neutralisation titre is predicted to reduce efficacy from 95% to 67% in a high efficacy vaccine, but from 70% to 25% for a vaccine with lower initial efficacy.

The analysis above investigates vaccine (and convalescent) protection against detectable SARS-CoV-2 infection (using the definitions provided in the different phase 3 and convalescent studies, see Supplementary Table 2). However, it is thought that the immune response may provide greater protection from severe infection than from mild infection. To investigate this, we also analysed data on the observed level of protection from severe infection where this was available (Supplementary Table 3, noting that the definition of severe infection was not consistent across studies). As there have been under 100 severe infections reported across all the phase 3 trials combined, the confidence intervals on the level of protection from severe infection are broad. The neutralisation level for 50% protection from severe infection was 3.1% of the average convalescent level (CI= 0.75% - 12.7%), which was significantly lower than the 20% level required for protection from any infection (p = 0.0004, likelihood ratio test, Supplementary Table 4). An important caveat to this analysis is the implicit assumption that neutralisation titre itself confers protection from severe infection, while cellular responses may also be important ^30-32^.

The estimated neutralisation level for protection from severe infection is approximately 6 times lower than the level required to protect from any infection. Thus, a higher level of protection against severe infection is expected for any given level of vaccine efficacy against mild (any) SARS-CoV-2 infection. Assuming that this relationship remains constant over time, it appears likely that immunity to severe infection may be much more durable than overall immunity to any infection. Long-term studies of antibody responses to other vaccines suggest that these responses generally stabilise with half-lives >10 years ^33,34^. Therefore we projected beyond the reported decay of SARS-CoV-2 responses (out to 8 months post-infection^5^) assuming that after 8 months post-infection the decay rate decreases exponentially (at rate 0.01d^-1^) until a 10-year half-life is achieved (details in Supplementary Methods). This analysis predicts that even without immune boosting a significant proportion of individuals may maintain protection from severe infection with an antigenically similar strain over the long term, even though they may become susceptible to mild infection (Figure 3B,C).

**Figure 3:**
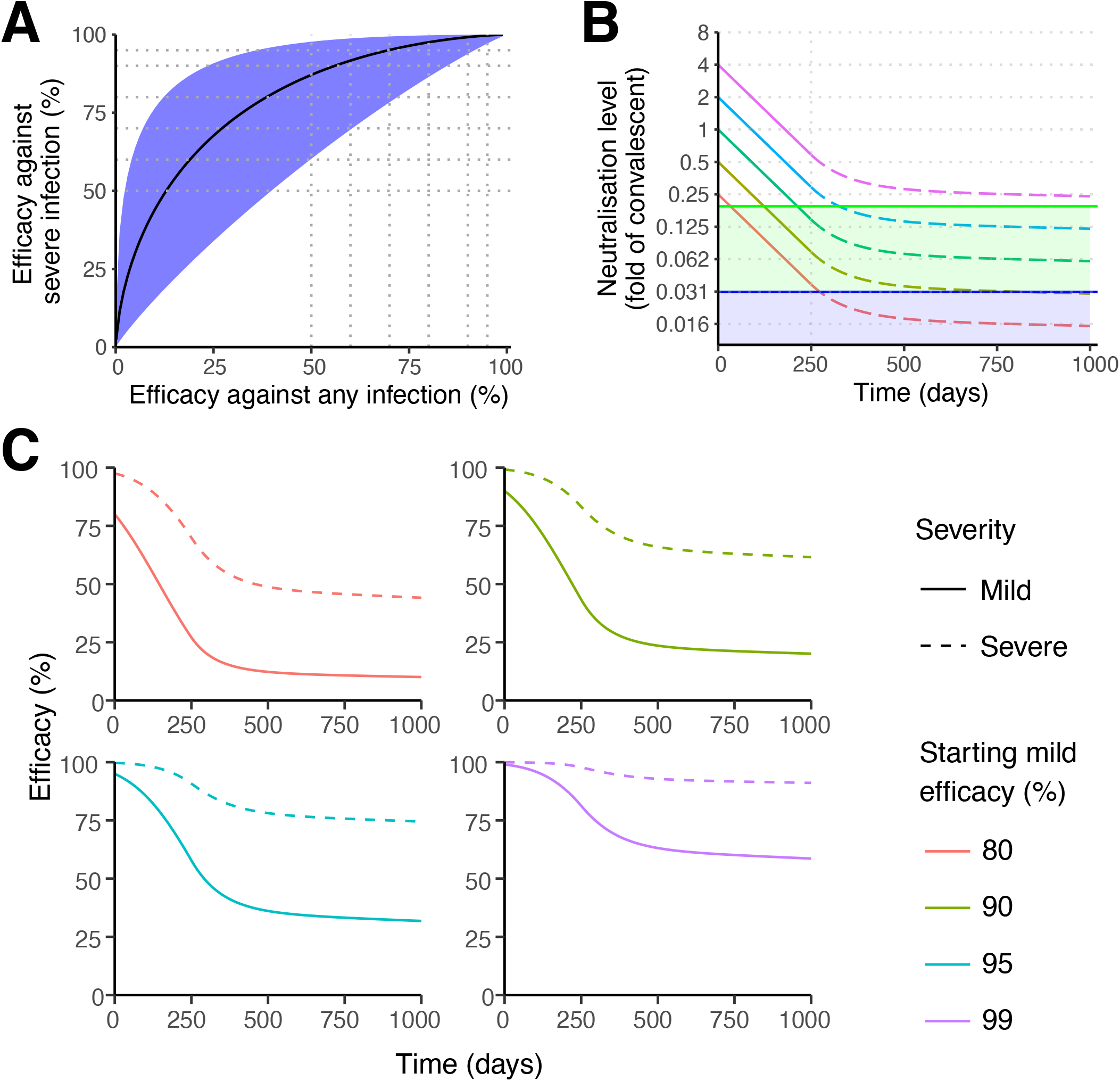
Protection from severe infection. A) The predicted relationship between efficacy against mild (any) SARS-CoV-2 infection (x-axis) versus efficacy against severe infection (y-axis). The black line indicates the best fit model for the relationship between protection against any versus severe SARS-CoV-2 infection. Shaded areas indicate 95% confidence intervals. Efficacy against severe infection was calculated by using a severe threshold that was a factor of 0.16 smaller than mild infection (CI = 0.039 to 0.66). B) Extrapolating the decay of neutralisation titres over time. This model assumes a half-life of SARS-CoV-2 neutralisation titre of 90 days over the first 250 days ^5^, after which the decay decreases (at rate 0.01d^-1^) until a 10-year half-life is achieved ^33,34^. For different initial starting levels the model projects the decay in level over the subsequent 1000 days. The green line indicates the predicted 50% protective titre from mild SARS-CoV-2 infection, and the purple line indicates the 50% protective titre from severe SARS-CoV-2 infection. The model illustrates that, depending on the initial neutralisation level, individuals may maintain protection from severe infection whilst becoming susceptible to mild infection (ie: with neutralisation levels remaining in the green shaded region). C) Extrapolating the trajectory of protection for groups with different starting levels of protection. The model uses the same assumptions on the rate of immune decay discussed in panel B. Note: The projections beyond 250 days rely on an assumption of how the decay in SARS-CoV-2 neutralisation titre will slow over time. In addition, the modelling only projects how decay in neutralisation is predicted to affect protection. Other mechanisms of immune protection may play important roles in providing long-term protection that are not captured in this simulation.

Understanding the relationship between measured immunity and clinical protection from SARS-CoV-2 infection is urgently needed for planning the next steps in the COVID-19 vaccine program. Placebo controlled vaccine studies are unlikely to be possible in the development of next generation vaccines, and therefore correlates of immunity will become increasingly important in planning booster doses of vaccine, prioritising next-generation vaccine development and powering efficacy studies ^35^. Our work uses available data on immune responses and protection to model both the protective titre and long-term behaviour of SARS-CoV-2 immunity. It suggests that neutralisation titre will be an important predictor of vaccine efficacy in future as new vaccines emerge. The model also predicts that immune protection from infection may wane with time as neutralisation levels decline and that booster immunisation may be required within a year. However, protection from severe infection may be considerably more durable, as lower levels of response may be required or alternative responses (such as cellular immune responses) may play a more prominent role. Our results are consistent with studies of both influenza and seasonal coronavirus infection, where reinfection is possible a year after initial infection, although usually resulting in mild infection^36,37^. Similarly, after influenza virus vaccination, protective efficacy is thought to decline by around 7% per month ^38^. Our modelling and predictions are based on a number of assumptions on the mechanisms and rate of loss of immunity. Important priorities for the field are the development of standardised assays to measure neutralisation and other immune responses, as well as standardised clinical trial protocols. These data will allow further testing and validation of other potential immune correlates of protection. However, our study develops a modelling framework for integrating available, if imperfect, data from vaccination and convalescent studies to provide a tool for predicting the uncertain future of SARS-CoV-2 immunity.

## Data Availability

All data is freely available upon request after publication.

## Ethics statement

This work was approved under the UNSW Sydney Human Research Ethics Committee (approval HC200242).

## Funding statement

This work is supported by an Australian government Medical Research Future Fund awards GNT2002073 (SJK, MPD, and AKW), MRF2005544 (to SJK, AKW, JAJ and MPD), MRF2005760 (to MPD), an NHMRC program grant GNT1149990 (SJK and MPD), the Victorian Government (SJK, AKW, JAJ), and Emergent Ventures Fast Grants (AWC). JAJ, DSK and SJK are supported by NHMRC fellowships. AKW, DC and MPD are supported by NHMRC Investigator grants. This study was supported by the Jack Ma Foundation (KS) and the A2 Milk Company (KS). The Melbourne WHO Collaborating Centre for Reference and Research on Influenza is supported by the Australian Government Department of Health.

## Notes

### Competing Interest Statement

The authors have declared no competing interest.

### Author Declarations

This work was approved under the UNSW Human Research Ethics Committee (approval HC200242).

