## Supplementary Methods and Figures for "What level of neutralising antibody protects from COVID-19?"

### Supplementary Methods and Results

#### Data extraction

Where possible, data values were used as directly stated in publications. In addition, where necessary, raw data was directly extracted from the original publications using an online digitizer tool (https://automeris.io/WebPlotDigitizer/).

#### Methods for Modelling and Data Analysis

##### Estimating the standard deviation of neutralisation titres

Neutralisation titres were extracted for each study (as above) and used to determine the standard deviation of the log-transformed neutralisation titres for each study. The standard deviation for each study had to consider that some measurements of neutralisation titre fell below the limit of detection (LOD) of that study assay (LOD varied for each study, table S1). To remove the effect of LOD censoring on estimates of the standard deviation of neutralisation titres we used a censored regression model to fit the distribution of the neutralisation titres for each study. The likelihood function is given by,

$$\mathcal{L}(\boldsymbol{D_s}|\mu_{cens},\sigma_{cens},L_s) = \prod_{n_i \in \boldsymbol{D_s}} f(n_i|\mu_{cens},\sigma_{cens})^{1-I_i} F(L_s|\mu_{cens},\sigma_{cens})^{I_i} \quad (1)$$

where, $\boldsymbol{D_s}$ is a vector of all the log-neutralisation titres, $n_i$, study $s$. The function $f$ is the probability function of a normal distribution with mean $\mu_{cens}$ and standard deviation $\sigma_{cens}$, and $F$ is the cumulative density function of the same distribution. The limit of detection of the assay for study $s$ is given by $L_s$ and when the index variable $I_i$ is 1 when $n_i \leq L_s$ and 0 otherwise. The negative log of this likelihood function was minimised in R using the built-in optimiser "nlm" to estimate the mean and standard deviation of the log transformed neutralisation titres after factoring in the LOD. When no limit of detection was reported or all values were above the LOD, the $L_s$ was set to $-Inf$.

*Pooled standard deviation*

Given the differing sample size of the neutralisation data for each study, the accuracy of estimations of the standard deviation for each study varied considerably. Therefore, despite finding some limited evidence of a differences in the standard deviation between each study (P=0.049, Fligner-Killeen test), we combined all extracted data and calculated a standard deviation of the pooled data. To do this we first centred the neutralisation data for each study at the reported mean of the neutralisation titres for that study. The limit of detection associated for each study was also adjusted in the same way. This provided a combined dataset of neutralisation titres from all studies, which was fitted using the likelihood model in equation 1 to the pooled data to produce an estimate of the standard deviation of the pooled data.

##### Modelling the relationship between neutralisation and protection.

In the above we used neutralisation titre information from each vaccine and convalescent individuals to estimate a distribution in neutralisation titres for each study. However, as discussed in the main text, given the diversity of assays used to assess neutralisation in each study (Table S1), from this point forward we normalise the neutralisation titres in each study by the mean of the neutralisation titre in the corresponding convalescent individuals contained within each study. Also, in all the analysis below we take the log-transform of the normalised neutralization titres. For simplicity, in the remainder of this document, we refer to these log-transformed normalised neutralisation titres as the **neutralisation levels**.

*Logistic method.*
To model the relationship between neutralisation level of antibodies collected from individuals after vaccination (or during convalescence) and protection from COVID-19 we assumed we assumed a logistic relationship between neutralisation level and protective efficacy, such that,

$$E_I(n \mid n_{50}, k) = \frac{1}{1 + e^{-k(n - n_{50})}} \tag{2}$$

where $E_I$ is the protective efficacy of an individual given the neutralisation level of $n$ (note the definition of neutralisation level above). The parameter $n_{50}$ is the neutralisation level at which an individual will have a 50% protective efficacy (i.e. half the chance of being infected compared with an unvaccinated person). The steepness of this relationship between protective efficacy and neutralisation level is determined by the parameter $k$.

We assume that a vaccine (or prior exposure) will induce a (normal) distribution of neutralisation levels ($n$) in a population with some mean $\mu_s$ and standard deviation $\sigma_s$. The mean neutralisation level for each study ($\mu_s$), is the difference between the log-transformed mean neutralisation titres for vaccinated and convalescent individuals within that study. The standard deviation ($\sigma_s$) is the standard deviation for vaccinated individuals in that study only. (Note that the distribution of neutralisation level for convalescent individuals has a mean of zero by definition (i.e. the log of the mean of neutralisation titres for convalescent individuals normalised by itself)). Therefore, the proportion of the vaccinated population for a study, $s$, that will be protected will be given by,

$$P(n_{50}, k, \mu_s, \sigma_s) = \int_{-\infty}^{\infty} E_I(n \mid n_{50}, k)\, f(n \mid \mu_s, \sigma_s)\ dn \tag{3}$$

where, $f$ is the probability density function of a normal distribution, and $E$ is the logisitic function in equation (2). The above integral is the so-called logistic-normal integral and the mean of the logit-normal distribution, which has no analytical solution[1]. Therefore, we use a simple numerical approximation (left Riemann sum).

*Fitting the logistic model and confidence intervals*
The above model of protection was fit to data on the protective efficacy of vaccines from Phase III (and another large cohort study of convalescent individuals). For each vaccine and convalescence study the number of: (1) control (unvaccinated/placebo/naïve) individuals enrolled ($N_s^c$), (2) control individuals infected ($I_s^c$), (3) vaccinated (previously exposed) individuals enrolled ($N_s^v$) and (4) vaccinated individuals infected ($I_s^v$) were used in fitting the model. The likelihood of observing the number of infected individuals in the control and vaccinated groups for each study, given some parameters, is,

$$\begin{aligned}\mathcal{L}_s(N_s^c, I_s^c, N_s^v, I_s^v, \mu_s, \sigma_s \mid n_{50}, k, b_s) \\ = Bi(N_s^c, I_s^c, b_s) Bi\big(N_s^v, I_s^v, b_s(1 - P(n_{50}, k, \mu_s, \sigma_s))\big)\end{aligned} \tag{4}$$

where $b_s$ is the probability of an unvaccinated control individual becoming infected in study $s$ (baseline risk), $b_s(1 - P(n_{50}, k, \mu_s, \sigma_s))$ is the probability of infection in the vaccination group (see eq. 3) and $Bi(N, K, p)$ is the binomial probability mass function of probability of $K$ events from a sample size of $N$, where each event has a probability $p$ of occurring. However, we wish to fit all studies simultaneously, and so the total likelihood of observing the data in all studies, given some parameters, is,

$$\mathcal{L}(\boldsymbol{N^c}, \boldsymbol{I^c}, \boldsymbol{N^v}, \boldsymbol{I^v}, \boldsymbol{\mu}, \boldsymbol{\sigma} \mid n_{50}, k, \boldsymbol{b}) = \prod_{\forall s} Bi(N_s^c, I_s^c, b_s) Bi\big(N_s^v, I_s^v, b_s(1 - P(n_{50}, k, \mu_s, \sigma_s))\big) \quad (5)$$

where $\boldsymbol{N^c}, \boldsymbol{I^c}, \boldsymbol{N^v}, \boldsymbol{I^v}, \boldsymbol{\mu}, \boldsymbol{\sigma}$ are vectors containing the data $N_s^c, I_s^c, N_s^v, I_s^v, \mu_s, \sigma_s$ for each study $s$, and $\boldsymbol{b}$ is a vector of the baseline risk parameters $b_s$ for each study. Best fitting parameters $n_{50}$, $k$ and $\boldsymbol{b}$ were found using the "nlm" optimiser in R by minimising $-\log(\mathcal{L})$. The standard error ($SE$) of these estimates were estimated using the hessian $H$ output from this function, and the formula $SE = \sqrt{\text{diag}(H^{-1})}$. The 95% confidence intervals were taken as $\pm 1.95 \times SE$ of the estimated parameters.

The variable $\mu_s$ is the mean neutralisation level, which can be calculated in two ways, firstly taking the geometric mean of neutralisation titres in vaccinated individuals over the geometric mean of the neutralisation titre in convalescent individuals in the same study. This is, in most cases, the ratio of two values directly reported in the immunogenicity studies. However, this approach does not account for situations in which the neutralisation assay had neutralisation titres below the limit of detection, so we also estimated this value by extracting the neutralisation titres from figures within each immunogenicity study (table S1), and computing the mean neutralisation titre for vaccinated and convalescent individuals in each study using censoring regression (equation (1)). Additionally, although it was in principle possible to compute the standard deviation of neutralisation levels for each study (as above), these appeared somewhat confounded by varying numbers of individuals between studies, hence we fit the above model using: 1) the standard deviation estimates for each study, 2) one standard deviation from one larger study to which we had direct access to raw data[2] (i.e. no manual data extraction required) and 3) an estimate of the standard deviation for all studies pooled together. The two different methods for estimating $\mu_s$ for each study, and the three methods for estimating $\sigma_s$ gives rise to 6 versions of the above model. All of these versions of the model were fitted, and the estimated protective levels were very similar (fig. S1).

*Protective Neutralisation Classification Model (PNC Model)*
The above modelling approach assumed neutralisation levels were normally distributed. Here we present a method for determining the protective threshold that is free of assumptions regarding the distribution of neutralisation levels. This model assumes that there is protective neutralisation level, $T$, above which individuals will be protected from infection and below which individuals will be susceptible. Using the protective efficacy observed in Phase III clinical trials of vaccinated individuals (and another large cohort study of convalescent individuals,[3]) (table S2), $E_s$, represents the proportion of individuals in each study who should possess a neutralisation level above the protective threshold. It follows then the number of individuals above the protective threshold in study $s$ is a function of $T$, which we denote $K_s(T)$. Therefore, the probability of observing $K_s(T)$ individuals above the protective threshold, given that there were $N_s$ individuals in the immunogenicity study (which are much smaller studies than the Phase III studies), is given by,

$$P(K_s(T) \mid N_s, T) = Bi(K_s(T), N_s, E_s) \quad (6)$$

where, $Bi$ is a binomial distribution. Note that,

$$K_s(T) = \sum_{n_i \in \boldsymbol{D_s}} H(n_i - T) \tag{7}$$

and $N_s = |\boldsymbol{D_s}|$, such that $H$ is the heavy-side step function taking the value 1 when $n_i - T > 0$, or 0 otherwise, and $|\boldsymbol{D_s}|$ denotes the size of set $\boldsymbol{D_s}$ (i.e. number of neutralisation levels measured). To determine one protective threshold using the results of all efficacy studies in this paper, we construct a likelihood function,

$$\mathcal{L}(\boldsymbol{D} | T) = \prod_{\boldsymbol{D_s} \in \boldsymbol{D}} Bi(K_s(T), N_s, E_s) \tag{8}$$

where $\boldsymbol{D}$ is the set of vectors of the neutralisation levels from each study. Note that this likelihood function is discontinuous as the threshold $T$ is varied. Therefore, we evaluate this likelihood function with the threshold $T$ set equal to all observed neutralisation levels $n_i$ across all studies, and find the $T$ value that maximises this likelihood (fig. S1). This method is determining a protective level at which the proportion of individuals with neutralisation levels above the threshold is in greatest agreement with the observed protective efficacy of that vaccine.

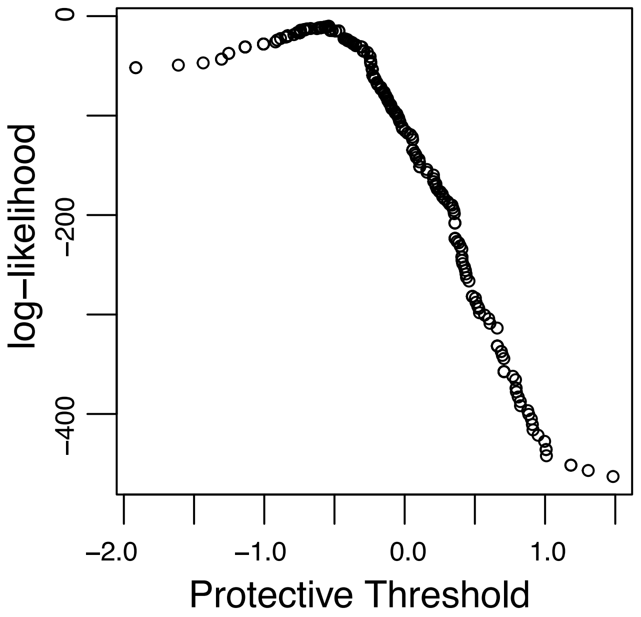


**Figure S1: Fitting the PNC Model.**
*This figure shows the estimated log-likelihood (log of equation 9) as a function of protective neutralisation threshold. To estimate the protective neutralisation threshold that is most consistent with the reported efficacy of each vaccine (and convalescence), the log-likelihood function (log of equation 9) was evaluated at all neutralisation levels observed in any of the studies listed in Supp Table 1, and the best fitting protective level was determined as the neutralisation level that maximised the likelihood function.*

Equation (8) is the likelihood function that should be adopted when neutralisation measurements are not impacted by a limit of detection. In the case that some neutralisation levels are measured below the limit of detection the likelihood function is adjusted as below,

$$\mathcal{L}(\boldsymbol{D}|\,T) = \prod_{\boldsymbol{D_s}\in\boldsymbol{D}} Bi(K_s(T), N_s, E_s)^{1-J_s} \times Q(C_s, N_s, 1-E_s)^{J_s} \tag{9}$$

where, $J_s$ is an index taking the value 1 when the LOD of study $s$ is above the threshold $T$ and at least one value is censored, or 0 otherwise. $C_s$ is the number of censored values in study $s$ and $Q$ is the cumulative binomial distribution function. This later term considers the probability that as many as all of the censored values were below the threshold T given the protective efficacy of the study $E_s$.

To determine the 95% confidence intervals for estimated protective neutralisation level a bootstrapping approach was used, in which the neutralisation levels were resampled 1000 times at random with replacement. Resampling was performed so as to preserve the total number of neutralisation levels in each study. These randomly generated samples of the original data were then fitted in the same way as described above, which generated 1000 corresponding estimates of the protective neutralisation level. The 95% confidence interval was calculated as the 2.5% and 97.5% percentiles of these 1000 estimates of the protective neutralisation level.

*Assessing predictive ability of model with a "Leave-one-out" analysis*
To determine the ability of the model to predict a vaccines efficacy, we performed a "leave one out" analysis, where we systematically excluded one of the vaccine (or convalescent) studies and performed the same model fitting procedure described above. Using the model fitted on the subset of the studies we estimated the efficacy of the vaccine that was "left out" from the fitted model. This "leave one out" analysis was performed with all versions of the logistic model (i.e. the 6 methods of estimating the mean neutralisation level and standard deviation outlined above were all considered). The predicted efficacy for each vaccine and convalescence obtained while leaving the study out are plotted against the reported efficacy in figure 1C. Also, the 50% protective level estimates each time a study was left out of the analysis provides a metric of the sensitivity of the model to the inclusion of each study. Note that excluding any of the studies did not greatly influence the estimate of the 50% protective level (fig. S2).

*Error bars and regions in efficacy and neutralisation*
In figure 1B there are both horizontal and vertical error bars as well a 95% predictive interval shown (shaded red) presented for the fitted model. The vertical errors bars indicate the 95% confidence intervals on the efficacy estimates for each study, these were calculated using the Katz-log method specified in Table 1 of[4]. The horizontal error bars indicate 95% confidence intervals in the difference of the mean of the ($\log_{10}$) neutralisation titres for vaccinated and convalescent individuals in each study. That is, these represent,

$$\pm 1.96 \times \sqrt{\frac{\sigma_v^2}{n_v} + \frac{\sigma_c^2}{n_c}}$$

where $\sigma_v$ and $\sigma_c$ are the standard deviation in the $\log_{10}$ of the neutralisation titres for vaccinated and convalescent individuals, respectively, and were estimated as described in section "*Estimating the standard deviation of neutralisation titres*" above. $n_v$ and $n_c$ are the number of vaccinated and convalescent individuals that were included for each study. The

95% predictive interval in figure 1 (shaded red region) was calculated using the delta method[5].

*Comparing the protective level in mild vs severe infection*

We also tested if the protective neutralisation level was different between mild and severe infection by fitting the combined dataset with two different mathematical models. The simplest model assumes we could use the same protective level in both severe and mild infection (ie, shared steepness parameter ($k$) and 50% protective level ($n_{50}$) in the logistic model (above)); and the alternative model uses different protective level parameters while we constrained the model to have the same $k$ in severe and mild infection (equation (3)). We used both the Akaike Information Criterion (AIC) and a likelihood ratio test to determine which model provided the best fit of the dataset for severe and all COVID-19 cases reported (Table S4).

**Modelling the decay of neutralisation and effects of antigenic variation.**

*Comparing decay in convalescence vs. vaccination.*

A number of studies have analysed the decay in neutralisation titre in convalescent subjects. These have generally shown a rapid early decay that slow with time[2,6-9]. We identified one study by Widge et. al.[10] where a time-course of neutralisation titre after mRNA vaccination was able to be analysed. This study measured decay of neutralisation titre out to 115 days. To compare the half-life of decay of neutralisation titre in vaccinated versus convalescent cohorts we analysed decay in this vaccination study compared with a previously published study of convalescent neutralisation titre[2], restricting the convalescent data over the same time intervals (fig. S3).

The Widge et al data[10] provided a timecourse of individual titres (as did the convalescent data), therefore we compared decay rates using linear mixed effects, by treating vaccination group as a binary variable. Statistical significance was determined based on the value of this covariate (if it was significantly different from zero), which was calculated by using the likelihood ratio test.

*Predicting the loss of efficacy as neutralisation wanes and due to variants*

The decay in efficacy with time (Figure 2A, B) was modelled by reducing the neutralisation levels at a rate corresponding to a half-life of 90 days and recalculating efficacy given this new distribution in neutralisation levels using equation (3). The efficacy against a SARS-CoV-2 variant, given an associated decrease in neutralisation (Figure 2C), was calculated by reducing the mean neutralisation level by a factor of 2, 5 or 10 and using equation (3). Efficacy against severe infection was similarly calculated using the same approach as above, but using the 50% protective level associated with the severe threshold (Table S4) that was a factor of 0.16 smaller than mild infection (CI = 0.039 to 0.66) (Figure 3C,E). We also extrapolated the decay of neutralisation level beyond the current data, assuming that neutralisation levels decay with a half-life of 90 days up until day 250, after which the decay decreases exponentially (at rate $0.01d^{-1}$) until a 10-year half-life is achieved (Figure 3D, E).

#### Results of Modelling

**Fitting with different models give similar estimates of protective neutralisation level**
Due to the limitations of reconciling data across the 7 vaccines (and studies of convalescent individuals) a number of approaches (described above) were employed to estimate the 50% protective neutralisation level. Additionally, variation in estimates of the 50% protective neutralisation level was explored when studies were excluded as a sensitivity analysis ("leave one out analysis"). All of the approaches applied produced similar estimates of the 50% protective level (fig. S2).

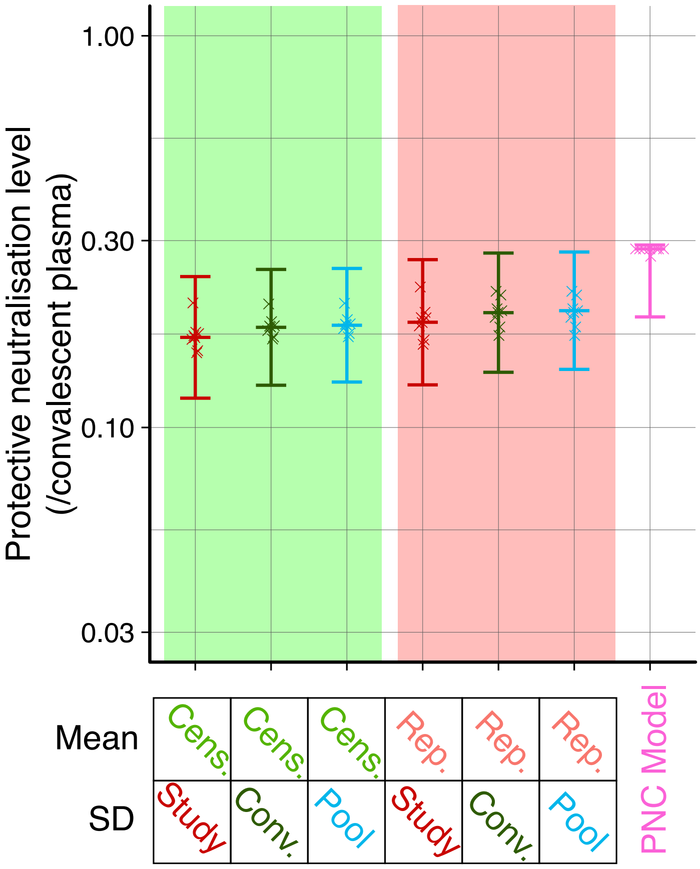


**Figure S2: Estimates of the 50% protective neutralisation level with different models.** *Two models, the logistic model (green and red shaded regions) and the protective neutralisation classification model (PNC Model, purple), were fit to the data on neutralisation levels in each study and protective efficacy of vaccination (and convalescence) from 8 studies. The logistic model was fit 6 times using a mean and standard deviation of the distribution in neutralisation levels for each study that was estimated under different methods described above. Central horizontal bars indicate the estimated 50% protective neutralisation level (for the logistic model), or protective threshold (for the PNC model) from each model. The error bars indicate the 95% confidence interval on these estimates. The 8 crosses overlayed on each model indicate the estimates of the protective level obtained when the same model is fit to only 7 of the 8 studies (i.e. excluding one study), and highlights that no single study strongly influences the estimate of protective neutralisation level.*
**Note that in this figure we present non-log transformed protective neutralisation levels to maintain consistency with the main text.*

**Estimated decay rates of neutralisation titres in vaccination and convalescence**

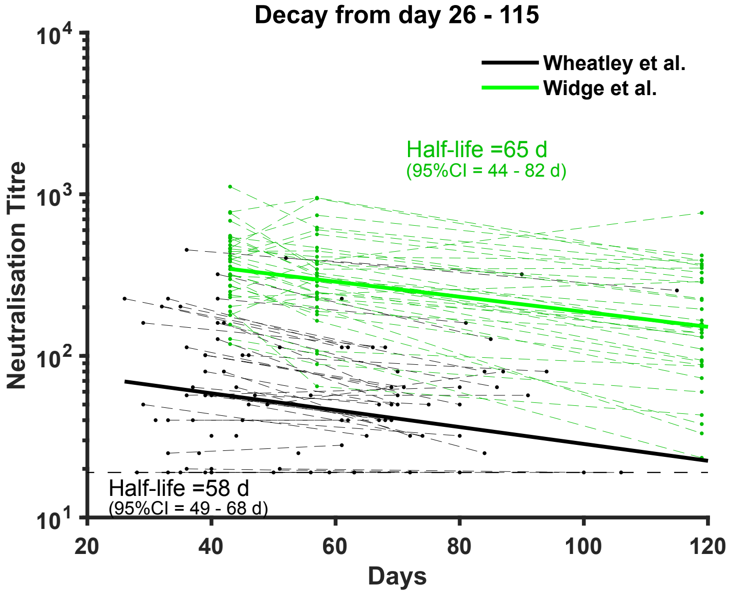


**Figure S3: Neutralisation titres reported in vaccined and convalescent individuals over time.**

*The decay half-lives estimated for a linear mixed effects model were not significantly different between vaccinated and convalescent individuals (P=0.88, likelihood ratio test).*

**Data sources**

| Manufacturer | Technical Name | Dosage | Dose 2 (Day) | Measured on Day | Assay* | Reference | Data derived from? |
|---|---|---|---|---|---|---|---|
| Moderna | mRNA-1273 | 100 µg | 29 | 42 | Live WT virus $PRNT_{80}$[1] | 11 | Table 2 |
| Novavax | NVX-CoV2373 | 5 µg + 50 µg Matrix-M1 adjuvant | 21 | 35 | Live WT MN $IC_{>99}$[2] | 12 | Figure 3, Panel B |
| Pfizer | BNT162b2 | 30 µg | 21 | 28 | Live fluorescent WT virus PRNT $IC_{50}$[3] | 13 | Figure 4, Panel B, combining data from all age groups |
| Sputnik | rAd26-S+rAd5-S | 10^11 viral particles | 21 | 42 | Live WT virus MN $IC_{50}$[4] | 14 | Figure 2, Panel D and Supp Figure 6-3 |
| Astrazeneca | ChAdOx1 nCoV-19 | 5x10^10 viral particles (SD) | 28 | 42 | Live WT virus MN $IC_{50}$[5] | 15 | Figure 4, Panel B |
| Johnson & Johnson | Ad26.COV2.S | 5x10^10 viral particles | N/A | 29 | Live WT virus PRNT $IC_{50}$[6] | 16 | Figure 2, Panel B, combining data from all age groups |
| Sinovac | CoronaVac | 3 µg | 14 | 28 | Live WT virus MN $IC_{50}$[7] | 17 | Figure 3, Panel C and supp fig 6-3 |

[1] Reduction in infectivity by at least 80% after 20 minutes incubation of serum dilution and SARS-CoV-2/human/USA/USA-WA1/2020 on Vero E6 cells, plaques counted at 3 days.

[2] WT virus microneutralisation assay using USA-WA1/2020 isolate on Vero E6 cells, counting vCPE at 72 hours. Inhibitory concentration of >99% deemed to be first sample dilution showing vCPE.

[3] Concentration yielding 50% reduction in fluorescent viral foci of genetically modified fluorescent virus (USA_WA1/2020 with mNeonGreen inserted into ORF7 of viral genome) on Vero CCL81 cells at 16-24 hours.

[4] WT Microneutralization assay with TCID50 of 100 (hCoV-19/Russia/Moscow_PMVL-1/2020). Cells and counting method not clear.

[5] Concentration giving absence of cytopathic effect (CPE) with 100 TCID50 of BavPat1/2020 isolate on Vero E6 cells, read at 4 days (described as 'Marburg virus neutralisation assay').

[6] 50% inhibitory concentration of 100 PFU Victoria/1/2020 strain virus on Vero E6 cells, serum incubated for 60-90 minutes, assay read at 24 hours.

[7] Microneutralisation assay – 100 CCID50 of the CHN/CN1/2020 strain incubated with serum dilution for 2 hours, vCPE read at 5 days (bespoke method for assigning inhibitory concentration from vCPE in duplicates, cell line not clear).

| | | | | | | | |
|---|---|---|---|---|---|---|---|
| Bharat Biotech | BBV152 | 6 µg with Algel-IMDG | 28 | 42 | Live WT virus $PRNT_{50}$[8] | [18] | Figure 2, Panel A |
| N/A | Convalescent Plasma | N/A | N/A | N/A | Live WT virus MN $IC_{50}$[9] | [2] | Figure 1, Panel B |

**Table S1: Data sources for Immunogenicity Data**

**WT = wild type, MN = micro-neutralisation, PRNT = plaque reduction neutralization test, MN = microneutralisation, IC = inhibitory concentration, numerical subscript = percentage neutralisation that is claimed in the assay.*

[8] Assay details in Supplementary Appendix, not yet available online.

[9] Microneutralisation using 100 TCID50 of CoV/Australia/VIC01/2020 incubated with serum dilution for one hour before placing on Vero cells. The neutralising antibody titre (IC50) was calculated 5 days post-inoculation using the Reed/Muench method.

| Manufacturer | Technical Name | Dosage | Dose 2 (Day) | Trial Started (Day) | Measure of effectiveness | Reference | Data derived from? |
|---|---|---|---|---|---|---|---|
| Moderna | mRNA-1273 | 100 µg | 29 | 42 | Prevention of primary symptomatic COVID-19 illness.[10] | [19] | Figure 4 |
| Novavax | NVX-CoV2373 | 5 µg + 50 µg Matrix-M1 adjuvant | 21 | 35 | Prevention of PCR-positive symptomatic mild, moderate, or severe COVID-19 illness[11]. | [20] | Text from press release |
| Pfizer | BNT162b2 | 30 µg | 21 | 28 | Prevention of primary covid-19 illness[12] | [21] | Table 2 |
| Sputnik | rAd26-S+rAd5-S | 10^11 viral particles | 21 | 21 | Prevention of PCR confirmed COVID-19 | [22] | Table 2 |
| Astrazeneca | ChAdOx1 nCoV-19 | 5x10^10 viral particles (SD) | 28 | 42 | Prevention of primary symptomatic COVID-19[13] | [23] | Table 2 (SD/SD only) |
| Johnson & Johnson | Ad26.COV2.S | 5x10^10 viral particles | N/A | 29 | Prevention of primary molecularly confirmed | [24] | Text from press release |

[10] Definition of symptomatic illness was: "Covid-19 cases were defined as occurring in participants who had at least two of the following symptoms: fever (temperature ≥38°C), chills, myalgia, headache, sore throat, or new olfactory or taste disorder, or as occurring in those who had at least one respiratory sign or symptom (including cough, shortness of breath, or clinical or radiographic evidence of pneumonia) and at least one nasopharyngeal swab, nasal swab, or saliva sample (or respiratory sample, if the participant was hospitalized) that was positive for SARS-CoV-2 by reverse-transcriptase–polymerase-chain-reaction (RT-PCR) test."

[11] Symptomatic disease defined as a minimum of either (i) fever (defined by subjective or objective measure, regardless of use of anti-pyretic medications), (ii) New onset cough or (iii) ≥ 2 of the following: (a) New onset or worsening of shortness of breath or difficulty breathing compared to baseline, (b) New onset fatigue, (c) New onset generalised muscle or body aches, (d) New onset headache lasting ≥ 48 hours, (e) New loss of taste or smell, (f) Acute onset of sore throat, congestion, and runny nose, (g) New onset nausea, vomiting, or diarrhea lasting ≥ 48 hours.

[12] Efficacy of BNT162b2 against confirmed Covid-19 with onset at least 7 days after the second dose in participants who had been without serologic or virologic evidence of SARS-CoV-2 infection up to 7 days after the second dose; …. Confirmed Covid-19 was defined according to the Food and Drug Administration (FDA) criteria as the presence of at least one of the following symptoms: fever, new or increased cough, new or increased shortness of breath, chills, new or in- creased muscle pain, new loss of taste or smell, sore throat, diarrhea, or vomiting, combined with a respiratory specimen obtained during the symptomatic period or within 4 days before or after it that was positive for SARS-CoV-2 by nucleic acid amplification–based testing, either at the central laboratory or at a local testing facility (using a protocol-defined acceptable test).

[13]Primary symptomatic covid-19 was defined as "virologically confirmed, symptomatic COVID-19, defined as a NAAT- positive swab combined with at least one qualifying symptom (fever ≥37·8°C, cough, shortness of breath, or anosmia or ageusia)."

| | | | | | | | |
|---|---|---|---|---|---|---|---|
| | | | | | moderate to severe/critical COVID-19 illness[14] | | |
| Sinovac | CoronaVac | 3 µg | 14 | Not reported | Prevention of COVID-19 | [25,26] | News / Media Articles |
| N/A | Convalescence | N/A | N/A | N/A | Prevention of PCR-Positive COVID-19 | [3] | Table 1 |

**Table S2: Data sources for Efficacy Data**

[14] Moderate defined as any one of the following new or worsening signs or symptoms (a) Respiratory rate ≥20 breaths/minute, (b)Abnormal saturation of oxygen (SpO2) but still >93% on room air at sea level, (c) Clinical or radiologic evidence of pneumonia, (d) Radiologic evidence of deep vein thrombosis (DVT), (e) Shortness of breath or difficulty breathing, or any two of the following new or worsening signs or symptoms: (i) Fever (≥38.0°C or ≥100.4°F), (ii) Heart rate ≥90 bpm, (iii) Shaking chills or rigors, (iv) Sore throat, (v) Cough, (vi) Malaise as evidenced by 1 or more of the following: Loss of appetite, Generally unwell, Fatigue, Physical weakness, (vii) Headache, (viii) Muscle pain (myalgia), (ix) Gastrointestinal symptoms (diarrhea, vomiting, nausea, abdominal pain) (x) New or changing olfactory or taste disorders, (xi) Red or bruised looking feet or toes.

| Manufacturer | Technical Name | Definition of Severe disease | Reference | Data derived from? |
|---|---|---|---|---|
| Moderna | mRNA-1273 | One of a list of severe criteria[15] | [19] | Text and Supplementary Table S16 |
| Novavax | NVX-CoV2373 | Participants meeting at least one of a list of endpoint criteria[16]. | [20] | Text from press release |
| Pfizer | BNT162b2 | Severe disease, occurring after the day of the first dose[17] | [21] | Supplementary Table S5 |
| Sputnik | rAd26-S+rAd5-S | Included moderate of severe disease (no further definition provided). | [22] | Table 2 |
| Astrazeneca | ChAdOx1 nCoV-19 | Includes both hospitalised and severe cases[18]. | [23] | Table 5 (all dose regimens) |
| Johnson & Johnson | Ad26.COV2.S | Prevention of primary molecularly confirmed moderate to severe/critical COVID-19 illness[19] | [24] | Text from press release, severe case numbers inferred to be the minimum numbers that could generate this efficacy. |

**Table S3: Data sources for data showing Efficacy against Severe Disease**

---

[15] Severe Covid-19 was defined as "one of the following criteria: respiratory rate of 30 or more breaths per minute; heart rate at or exceeding 125 beats per minute; oxygen saturation at 93% or less while the participant was breathing ambient air at sea level or a ratio of the partial pressure of oxygen to the fraction of inspired oxygen below 300 mm Hg; respiratory failure; acute respiratory distress syndrome; evidence of shock (systolic blood pressure <90 mm Hg, diastolic blood pressure <60 mm Hg, or a need for vasopressors); clinically significant acute renal, hepatic, or neurologic dysfunction; admission to an intensive care unit; or death.

[16] Endpoint criteria for severe disease were listed as: (a) Tachypnea: ≥ 30 breaths per minute at rest, (b) Resting heart rate ≥ 125 beats per minute, (c) SpO2: ≤ 93% on room air or PAO2/FiO2 < 300, (d) High flow oxygen therapy or NIV/NIPPV (e.g., CPAP or BiPAP) , (e) Mechanical ventilation or ECMO, (f) One or more major organ system dysfunction or failure (e.g., cardiac/circulatory, pulmonary, renal, hepatic, and/or neurological, to be defined by diagnostic testing/clinical syndrome/interventions), including any of the following: i. ARDS, ii. Acute renal failure, iii. Acute hepatic failure, iv. Acute right or left heart failure, v. Septic or cardiogenic shock (with shock defined as SBP < 90 mm Hg OR DBP < 60 mm Hg, vi. Acute stroke (ischemic or hemorrhagic), vii. Acute thrombotic event: AMI, DVT, PE, viii. Requirement for: vasopressors, systemic corticosteroids, or hemodialysis, (g) Admission to an ICU, (h) Death

[17] Severe Covid-19 is defined as confirmed Covid-19 with one of the following additional features: clinical signs at rest that are indicative of severe systemic illness; respiratory failure; evidence of shock; significant acute renal, hepatic, or neurologic dysfunction; admission to an in- tensive care unit; or death.

[18] Hospitalised cases have WHO clinical progression score ≥ 4. Severe cases were defined as those with WHO clinical progression score ≥ 6.

[19] Severe disease defined as one or more of the following: (a) Clinical signs at rest indicative of severe systemic illness (respiratory rate ≥30 breaths/minute, heart rate ≥125 beats/minute, oxygen saturation (SpO2) ≤93% on room air at sea level, or partial pressure of oxygen/fraction of inspired oxygen (PaO2/FiO2) <300 mmHg), (b) Respiratory failure (defined as needing high-flow oxygen, non-invasive ventilation, mechanical ventilation, or extracorporeal membrane oxygenation [ECMO]), (c) Evidence of shock (defined as systolic blood pressure <90 mmHg, diastolic blood pressure<60 mmHg, or requiring vasopressors), (d) Significant acute renal, hepatic, or neurologic dysfunction, (e)Admission to the ICU, (f) Death

| **Model Structure** | **LL** | **AIC** | **Slope ($k$)** | **Severe $n_{50}$** | **Mild $n_{50}$** | **p-value** |
|---|---|---|---|---|---|---|
| **Different $n_{50}$** | -66.22 | 138.44 | 3.31(2.22 – 4.95) | 0.03 (0.0075 – 0.13) | 0.19 (0.14 – 0.27) | 0.0004 |
| **Same $n_{50}$** | -72.48 | 148.96 | 3.64 (2.4 – 5.5) | 0.19 (0.14 – 0.26) | 0.19 (0.14 – 0.26) | |

**Table S4: Model comparison of the protective level in mild and severe infection**

*Table shows log-likelihood, AIC and parameter estimates (95% CI), for a logistic model having either the same (top row) or different (bottom row) 50% protective level ($n_{50}$) for severe and mild infection. p-value was obtained from a likelihood ratio test comparing the two models.*
